# Recurrent SARS-CoV-2 RNA positivity after COVID-19: A systematic review and meta-analysis

**DOI:** 10.1101/2020.07.19.20157453

**Authors:** Mahalul Azam, Rina Sulistiana, Martha Ratnawati, Arulita Ika Fibriana, Udin Bahrudin, Syed Mohamed Aljunid

## Abstract

**Background:** Previous studies reported recurrent SARS-CoV-2 RNA positivity in individuals who had recovered from COVID-19 infections. However, little is known regarding the systematic review of recurrent SARS-CoV-2 RNA positivity. The current study conducted a systematic review and meta-analysis, aimed to estimate the incidence of recurrent SARS-CoV-2 RNA positivity after recovery from COVID-19 and to determine the factors associated with recurrent positivity.

**Methods:** We searched the PubMed, MedRxiv, BioRxiv, the Cochrane Library, ClinicalTrials.gov, and the World Health Organization International Clinical Trials Registry for studies published to June 12, 2020. Studies were reviewed to determine the risk of bias. A random-effects model was used to pool results. Heterogeneity was assessed using *I*^*2*^.

**Results:** Fourteen studies of 2,568 individuals were included. The incidence of recurrent SARS-CoV-2 positivity was 14.81% (95% confidence interval [CI]: 11.44–18.19%). The pooled estimate of the interval from disease onset to recurrence was 35.44 days (95% CI: 32.65–38.24 days), and from the last negative to recurrent positive result was 9.76 days (95% CI: 7.31–12.22 days). Patients with younger age (mean difference [MD]=-2.27, 95% CI: -2.95 to -1.80) and a longer initial illness (MD=8.24 days; 95% CI: 7.54 – 8.95; *I*^*2*^=98.9%) were more likely to experience recurrent SARS-CoV-2 positivity, while patients with diabetes (RR=0.52; 95% CI: 0.30-0.90; *I*^*2*^=53%), severe disease (RR=0.54; 95% CI: 0.35-0.84; *I*^*2*^=70%), and a low lymphocyte count (RR=0.58; 95% CI: 0.39 – 0.86; *I*^*2*^=48%) were less likely to experience recurrent SARS-CoV-2 positivity.

**Conclusions:** The incidence of recurrent SARS-CoV-2 positivity was 14.81%. The estimated interval from disease onset to repeat positivity was 35.44 days, and the estimated interval from the last negative result to recurrent positive result duration was 9.76 days.

## Background

Globally, the reported number of confirmed infections and deaths due to the severe acute respiratory syndrome coronavirus 2 (SARS-CoV-2 pandemic) was 7,410,510 and 418,294, respectively, by June 12 2020.[1] Country governments have implemented public health measures such as lockdowns, physical distancing, use of face masks, and frequent hand-washing; however, the incidence of SARS-CoV-2 infection is still increasing. The proportion of severe cases and case fatality rates have been reported to be 25.6% and 3.6%, respectively,[2] with individuals with comorbidities being at greater risk of developing severe disease.[2,3]

The World Health Organization (WHO) has provided criteria for assessing the recovery of patients hospitalized with coronavirus disease 2019 (COVID-19).[4] Recently, there have been several reports of recurrent SARS-CoV-2 RNA positivity in individuals who had recovered from COVID-19.[5,6] It was estimated that the incidence of recurrent SARS-CoV-2 positivity in individuals who have recovered from COVID-19, ranged from 7.3%[5] to 21.4%.[6] However, to date no systematic reviews have been published to provide a pooled estimate of the incidence of recurrent positivity.. This systematic review aimed to: estimate the incidence of recurrent SARS-CoV-2 positivity and determine the characteristics and risk factors related to the recurrent SARS-CoV-2 positivity in patients who had recovered from COVID-19.

## Methods

### Protocol and registration

This review is written following the Preferred Reporting Items for Systematic Review, and Meta-Analysis Protocol (PRISMA-P).[7] The protocol of this review was published in the International Prospective Register of Systematic Reviews (PROSPERO) on May 14, 2020, reference no. CRD42020186306.[8]

### Search strategy and information resources

A search was conducted on PubMed, MedRxiv, BioRxiv, Cochrane Library, ClinicalTrials.gov, the WHO international register of clinical trials registry using the search term in Medical Subjects Headings (MeSH) and free text: (“2019 nCoV” OR “2019nCoV” OR “2019 novel coronavirus” OR “COVID 19” OR “COVID19” OR “new coronavirus” OR “novel coronavirus” OR “SARS CoV-2” OR (Wuhan AND coronavirus) OR “COVID 19” OR “SARS-CoV” OR “2019-nCoV” OR “SARS-CoV-2”) AND ((recurrence) OR (relapse) OR (re*infection) OR (re*activation)).

### Data management and study selection

Literature search results were organized using Mendeley (Mendeley, Ltd, Elsevier, UK). Article titles and abstracts retrieved from the databases were transferred to Mendeley citation manager after being screened and checked for duplication. All records that did not meet the eligibility criteria were excluded from the review.

The eligibility of articles based on their title and abstract was assessed independently by MA and AF. If necessary, the full paper was retrieved to further determine the eligibility status. In cases of disagreement regarding eligibility, consensus was reached by consulting a third reviewer (MR). The eligibility criteria were: (i) the study designs are cross-sectional, case-control or cohort design; (ii) the study reports the incidence of recurrent SARS-CoV-2 positivity in individuals who had recovered from COVID-19 and its related factors; and (iii) the articles included published or unpublished studies. The published studies may included both peer-reviewed reports and pre-print reports. Studies in languages other than English were excluded if no translated version of the manuscript was available.

The full text of the articles that met the eligibility for the review were then assessed. If the data provided in the articles were incomplete, the author was contacted to obtain complete data. Data collection forms were used for specific purposes, including the screening process, determining eligibility, data collection, and incomplete data identification as well as the risk of bias assessment.

### Data extraction and quality assessment

The following data items were extracted: authors, funding, study design, the population of the study, number of episodes of recurrent SARS-CoV-2 positivity per case, and patient characteristics. The patient characteristics considered included age, sex, body mass index, clinical/laboratory manifestations, and comorbidities such as diabetes and hypertension. The outcome was recurrent SARS-CoV-2 positivity in individuals who had recovered from COVID-19, determined as based on positive result of reverse transcription polymerase chain reaction (RT-PCR)on re-testing, after being followed-up or re-admitted after discharged from hospital.

We used the quality assessment tool for cross-sectional and cohort studies published by the National Institutes of Health to assess the methodological quality of included studies.[9] Each item was scored 0 or 1 point based on the criteria. A total of all items ranged from 0 to 14 was used to assess the quality of the article. Based on the overall score, we categorized articles to high risk of bias with score ≤6, medium risk of bias with a score of 7–10, and low risk of bias when the score was ≥11. Each study was assessed for risk of bias independently by MA and MR. Any disagreement in the risk of bias assessment was resolved by discussion to reach consensus or by consulting UB and SA.

### Data analysis

We performed data analysis using Revman (Review Manager version 5.3.5 Copenhagen, The Nordic Cochrane Centre, 2014). Random-effects meta-analysis was used to calculate the pooled incidence of recurrent SARS-CoV-2 positivity with 95% confidence intervals. The incidence for each individual study with its standard error (SE) adds to the study data in RevMan. If the SE was not reported and the raw data could not be accessed, the SE was calculated using the formula 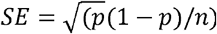. Meta-analysis was used to calculate pooled estimates of the time from disease onset to recurrent test positivity and the time from the last negative test result to recurrent positivity.

Meta-analysis was also used to calculate the pooled relative risk (RR) of recurrent SARS-CoV-2 positivity according to age, sex, hypertension, diabetes, other co-morbidities, disease severity, body mass index (BMI), fever as the initial presenting compliant, days from onset to negative conversion, lymphocyte count, D-dimer, and lung consolidation. We then assessed the heterogeneity between studies using *I*^*2*^, with values of 25%, 50%, and 75% representing low, moderate, and high heterogeneity, respectively.

## Results

Our search on June 12, 2020, produced 397 records. Of these records, 392 were left after the duplicates were removed. Of these records, 371 were excluded from the review because the articles did not report recurrent SARS-CoV-2 positivity. Of this, 21 full texts were assessed for eligibility and 14 studies were included in the meta-analysis (Figure 1).

**Figure 1.**
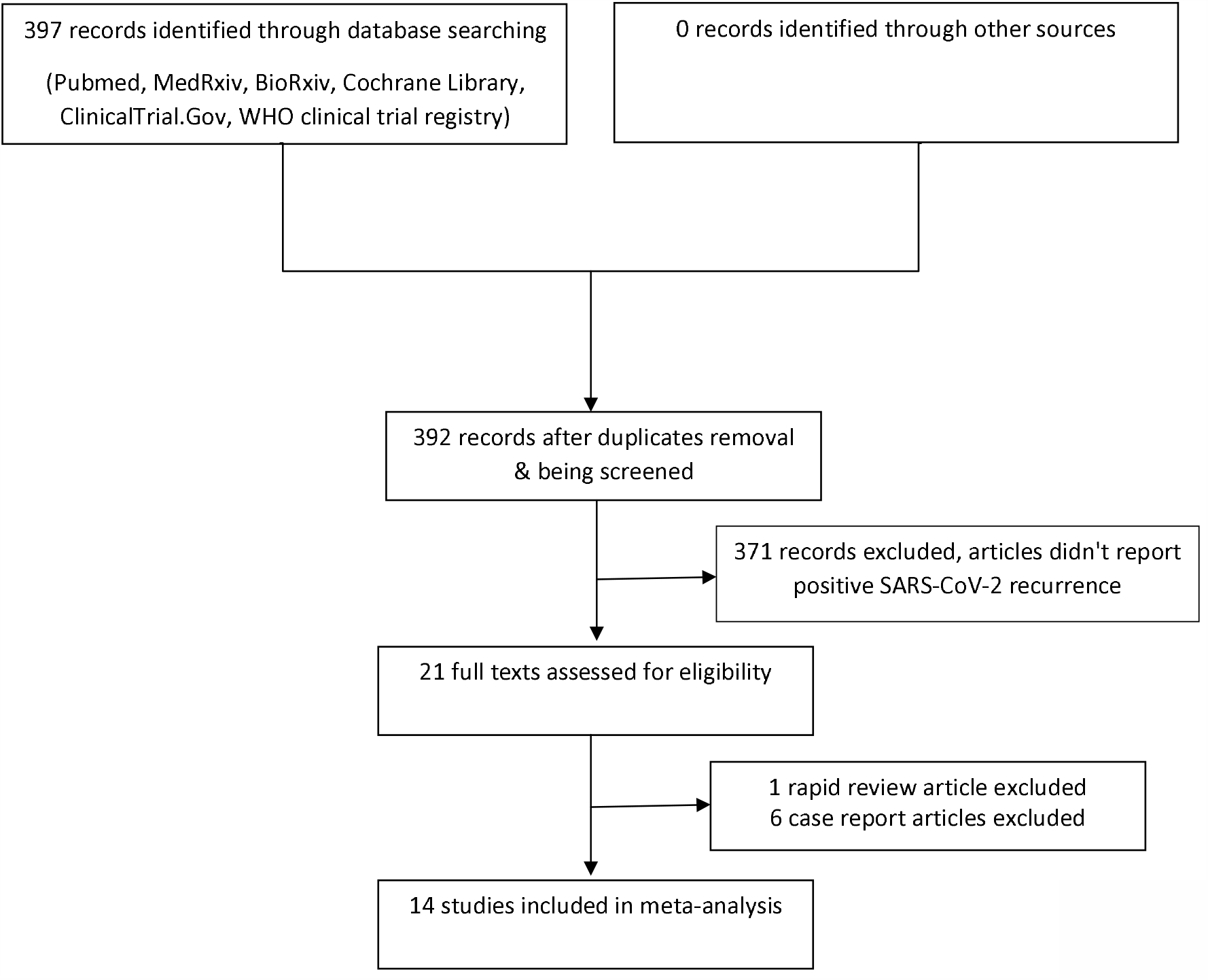
PRISMA-P study selection diagram.

Table 1 summarizes the characteristics of the finally selected studies. These studies were published between Mar 17 and May 29, 2020. We included four non-peer-reviewed studies. There was a total of 2,568 participants from all the studies combined, of which 318 experienced recurrent SARS-CoV-2 positivity. Thirteen of the 14 studies were conducted in China and one study was conducted in Brunei. Four of the Chinese studies were conducted in the city of Wuhan and the rest were conducted in other cities. There were six studies (43%) with a cross-sectional design, and four studies (29%) each with a retrospective cohort and prospective cohort design. The most frequently used sample types were nasopharyngeal or oropharyngeal swabs alone (46%), and the remaining studies used a variety of sample types including fecal, nasopharyngeal, and oropharyngeal swabs. One study did not report the type of sample that was used.[10] The median interval duration from disease onset to recurrence ranged from 21 to 50 days, while the interval from the last negative to recurrent positive result ranged from 4 to 19 days. The risk of bias was assessed as low in seven studies (50%), moderate in six studies (43%), and high in one study (7%) (Supplementary Table 1).

**Table 1.**
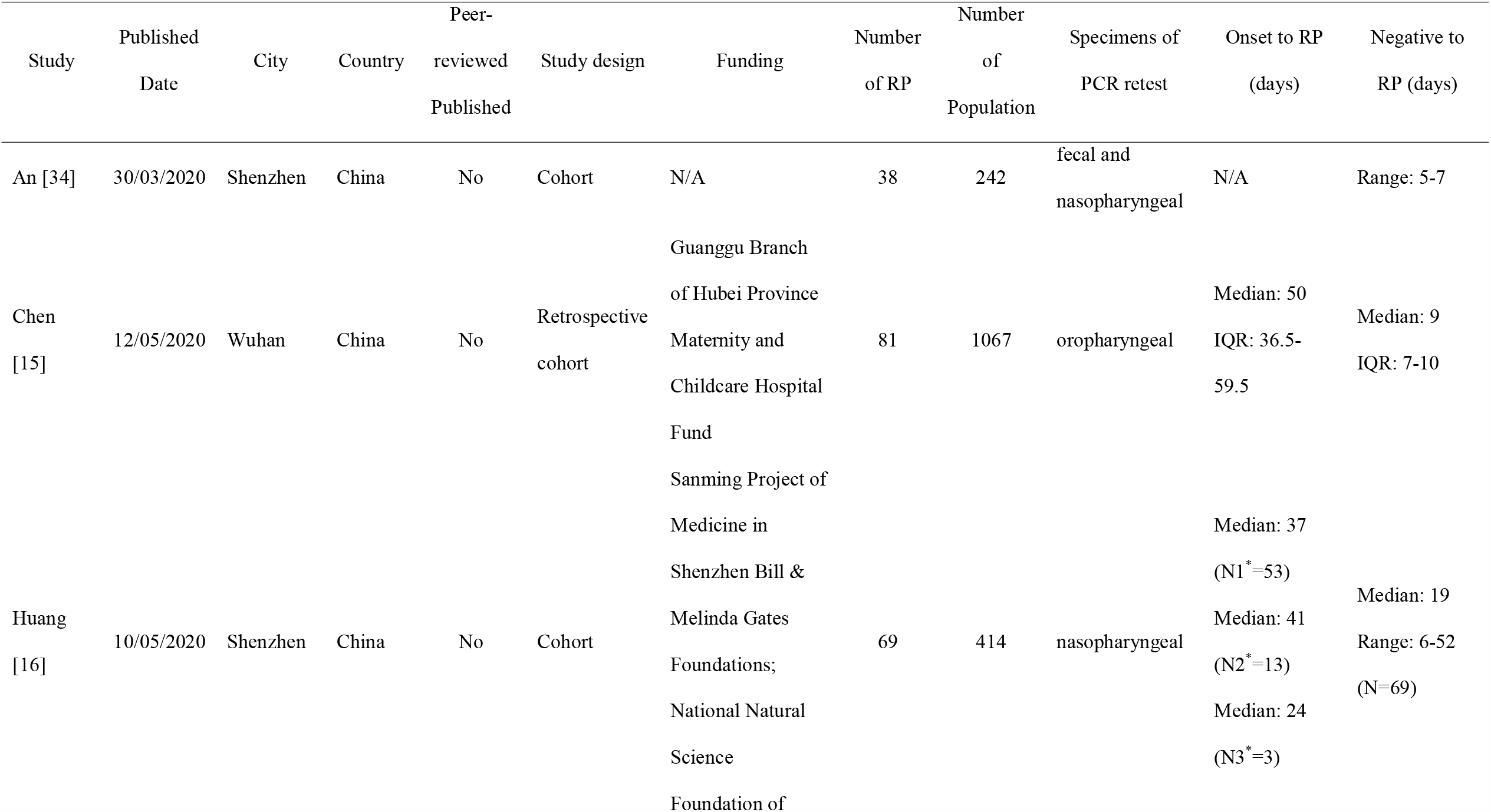
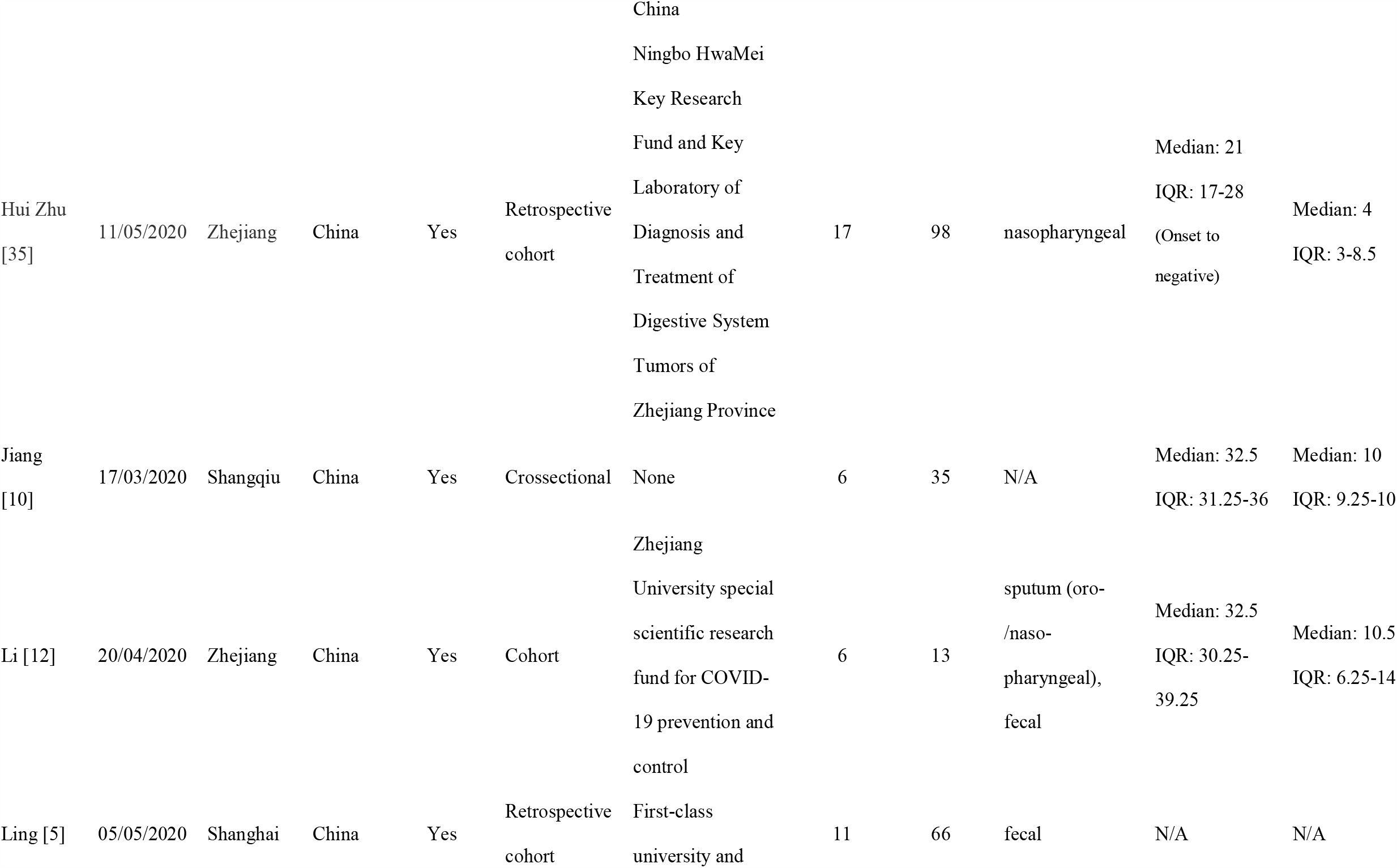
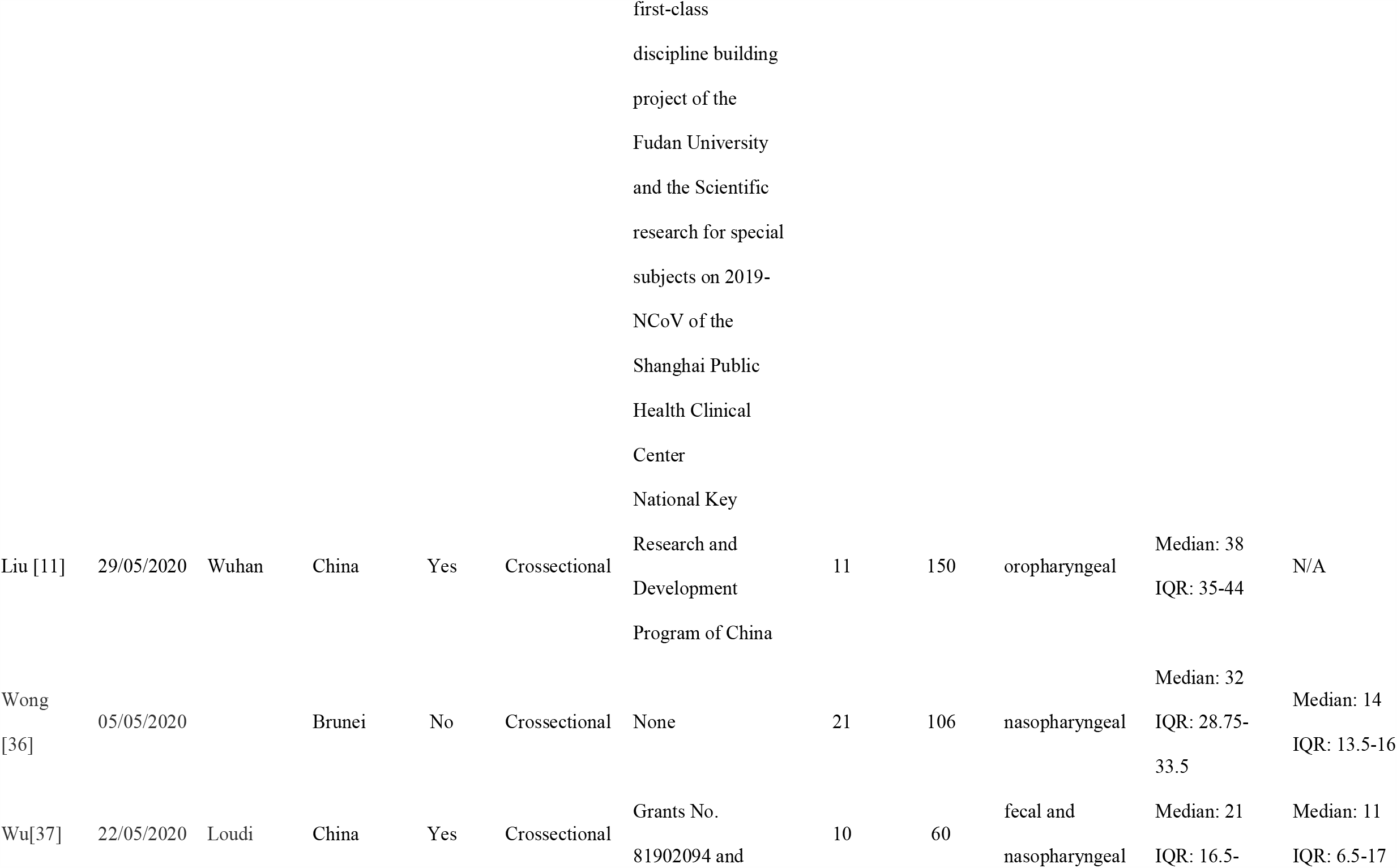
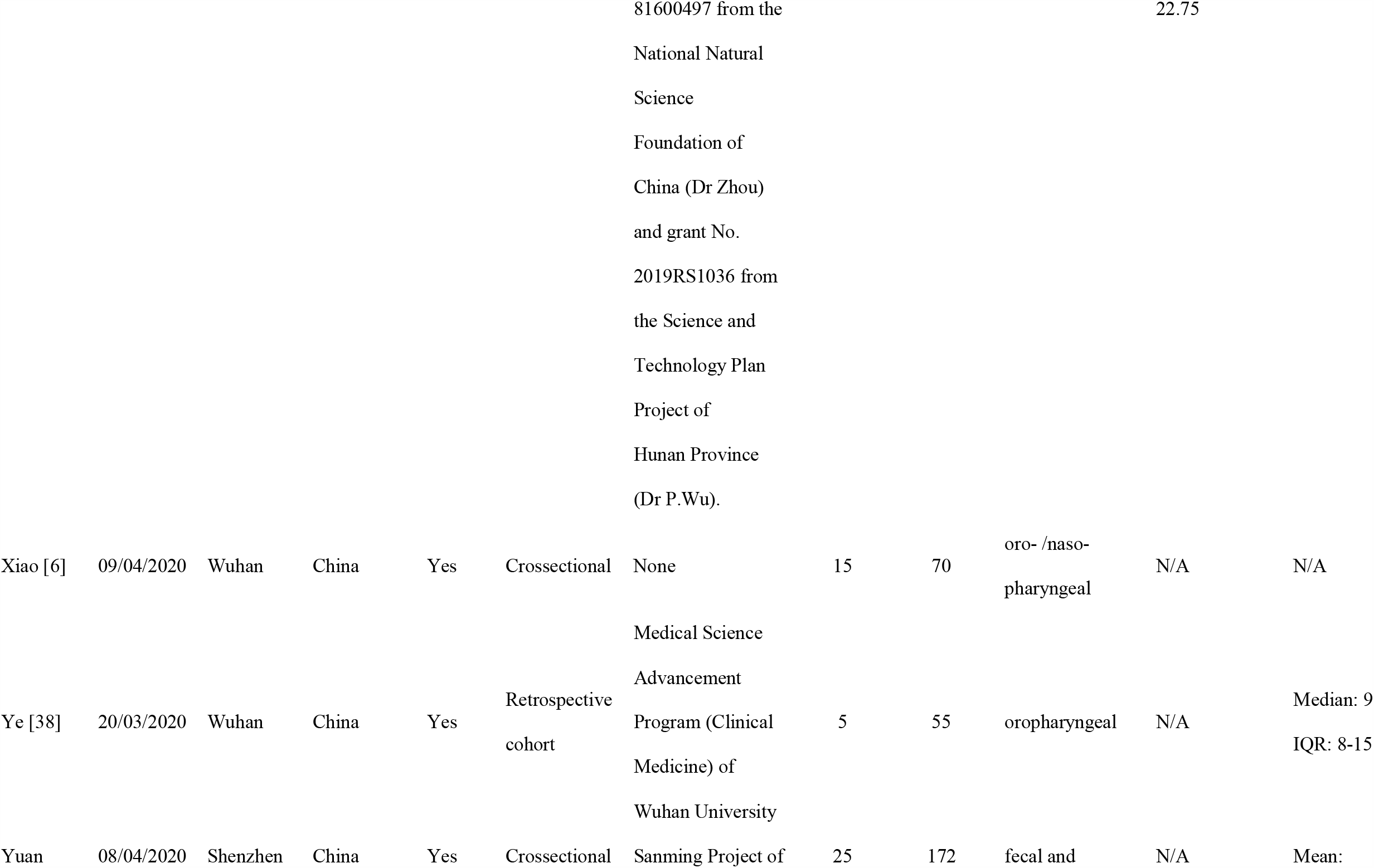
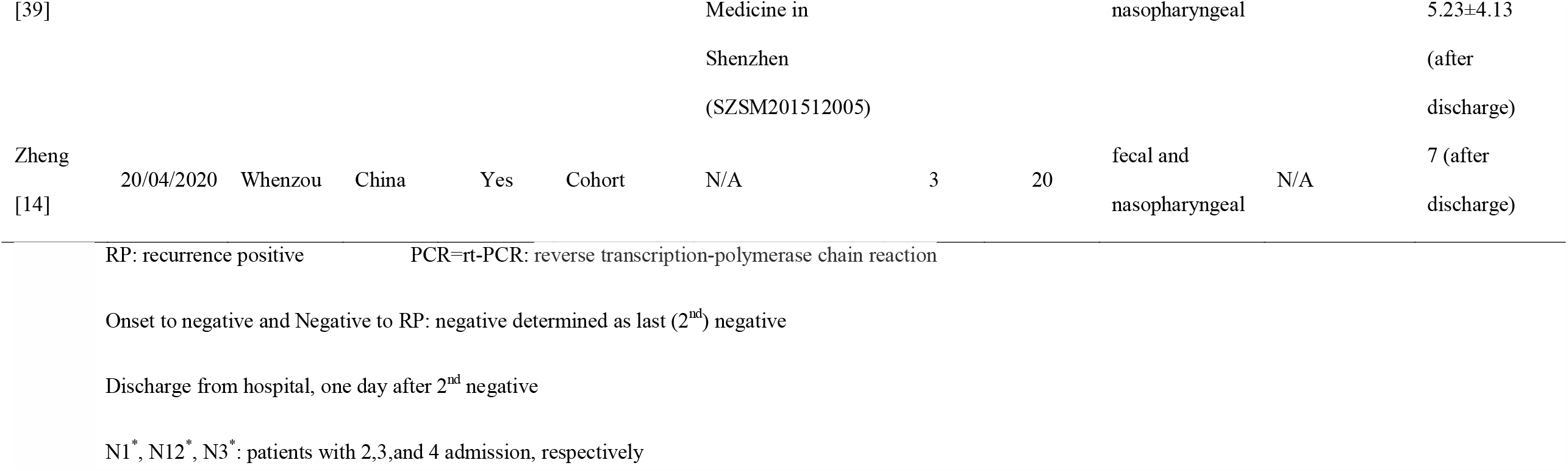
Study characteristics included in the meta-analysis

The pooled estimate of the incidence of recurrent SARS-CoV-2 positivity was 14.81% (95% CI: 11.44–18.19%) (Figure 2). Liu et al.[11] found the lowest incidence (7.33%, N=150), and Li et al.[12] found the highest incidence (46.2%, N=13).

**Figure 2.**
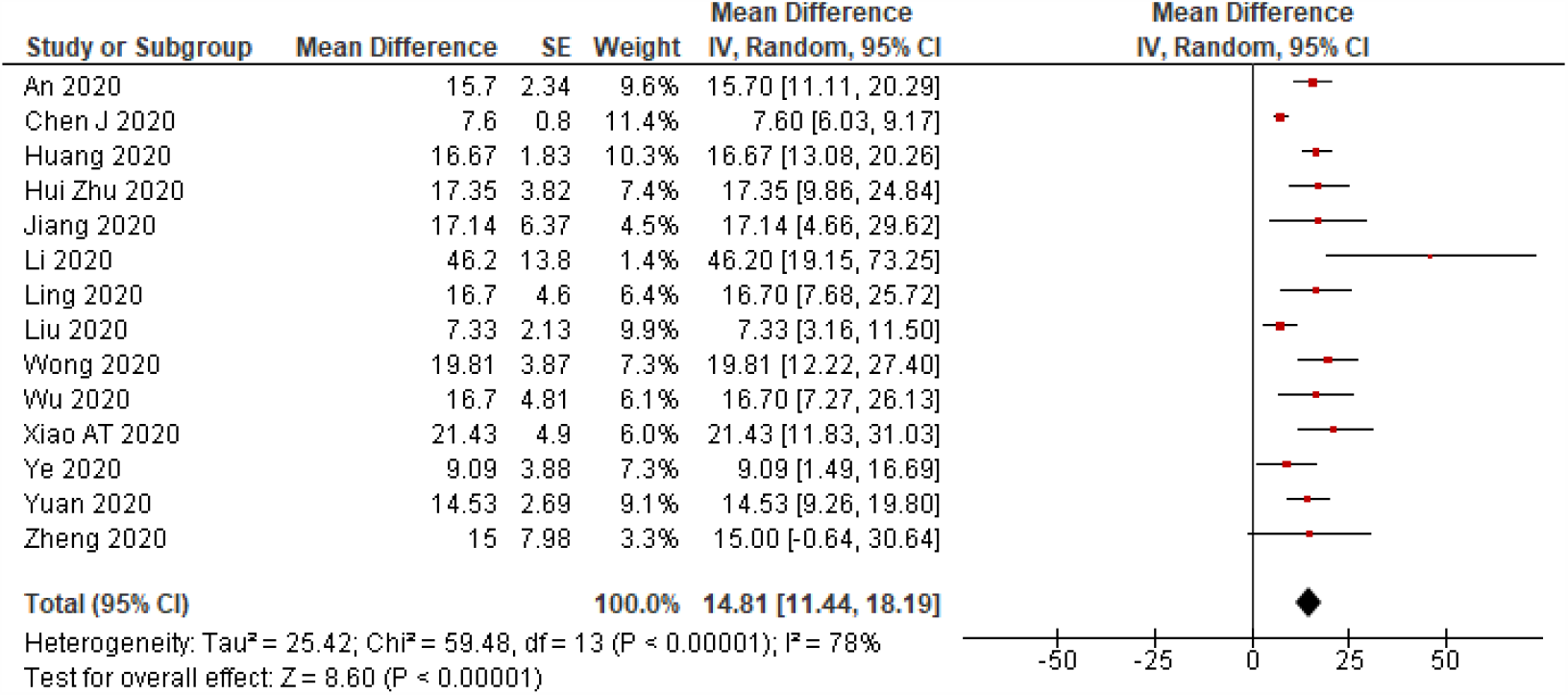
A meta-analysis of the pooled estimated incidence of SARS-CoV-2 RNA positivity.

Seven studies provided results on the time from disease onset to recurrent positivity, and eight studies provided results on the time from testing negative to recurrent positivity. The pooled estimate of the interval from disease onset to recurrent positivity was 35.44 days (95% CI: 32.65–38.24 days), and the pooled estimate of the last negative to recurrent positivity was 9.76 days (95% CI: 7.31–12.22 days) (Figures 3 and 4).

**Figure 3.**
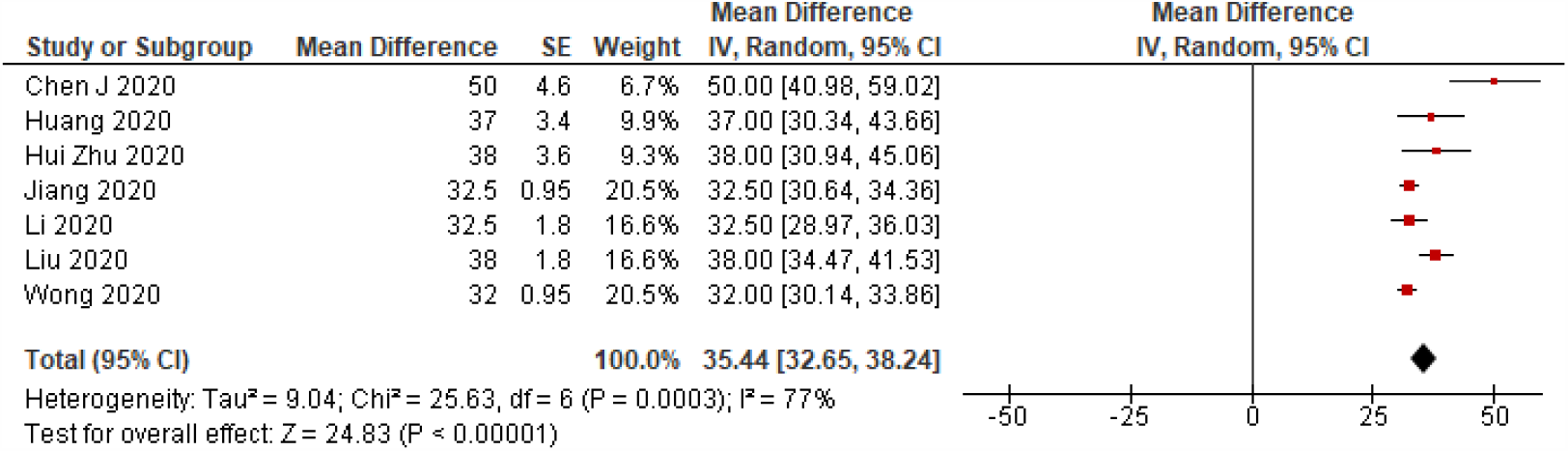
A meta-analysis of the pooled estimated onset to recurrent SARS-CoV-2 RNA positivity (days)

**Figure 4.**
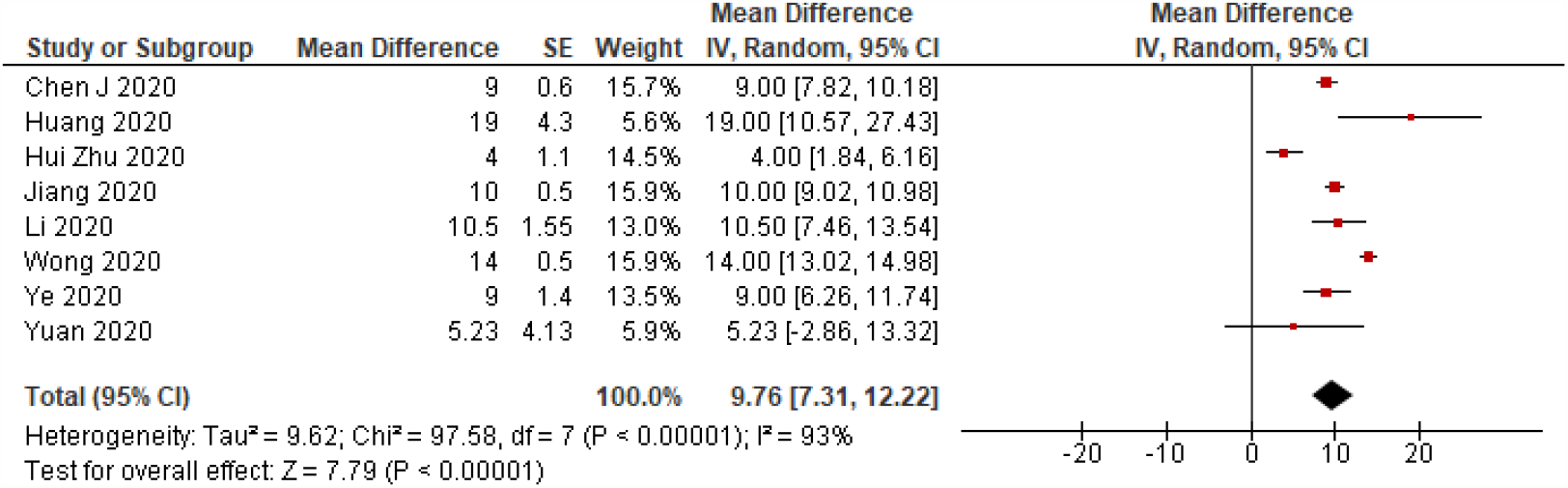
A meta-analysis of the pooled estimated last negative to recurrent SARS-CoV-2 RNA positivity (days)

Patients with younger age were more likely to experience recurrent SARS-CoV-2 positivity (mean difference: −2.27, 95% CI: −2.95 to −1.80), but there was considerable heterogeneity between studies in the effect of age (*I*^*2*^=99%). Patients with diabetes were less likely to experience recurrent SARS-CoV-2 positivity (RR: 0.52, 95% CI: 0.30-0.90, *I*^*2*^=53%). Patients with severe COVID-19 were also less likely to experience recurrently positivity than those with less severe disease (RR: 0.54, 95% CI: 0.35-0.84, *I*^*2*^=70%). A longer interval from disease onset to the last negative PCR result during the first admission was associated with a greater risk of recurrent SARS-CoV-2 positivity (mean difference: 8.24 days, 95% CI: 7.54–8.95 days, *I*^*2*^=98.9%). Patients with a low lymphocyte count (<1.1×10^9^/L) had a higher risk of experiencing recurrent SARS-CoV-2 positivity (RR: 0.58, 95% CI: 0.39–0.86, *I*^*2*^=48%). We did not find an association between sex, BMI, co-morbidity, hypertension, fever, lung consolidation, or D-dimer and the risk of recurrent SARS-CoV-2 positivity (Figure 5).

**Figure 5.**
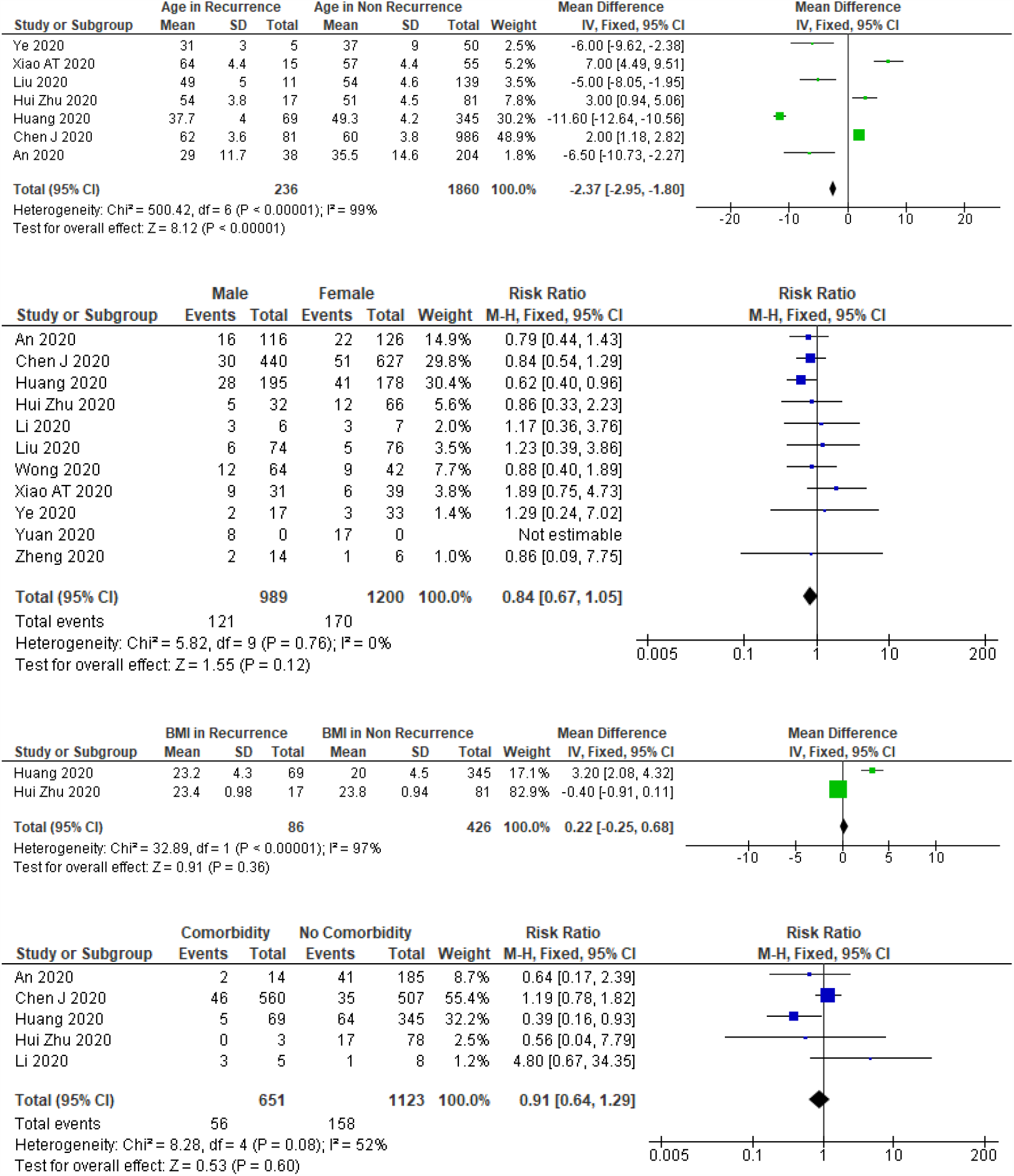
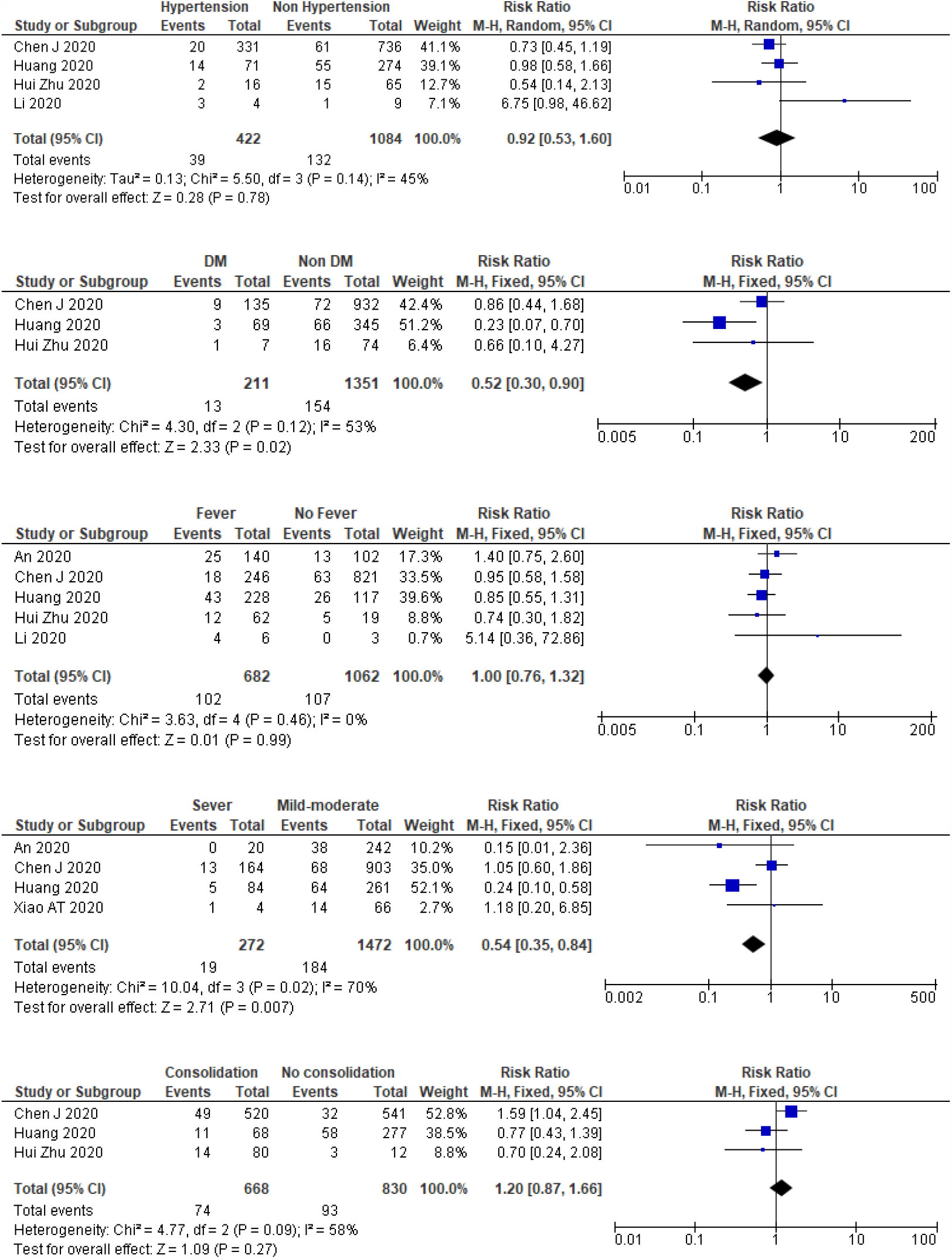
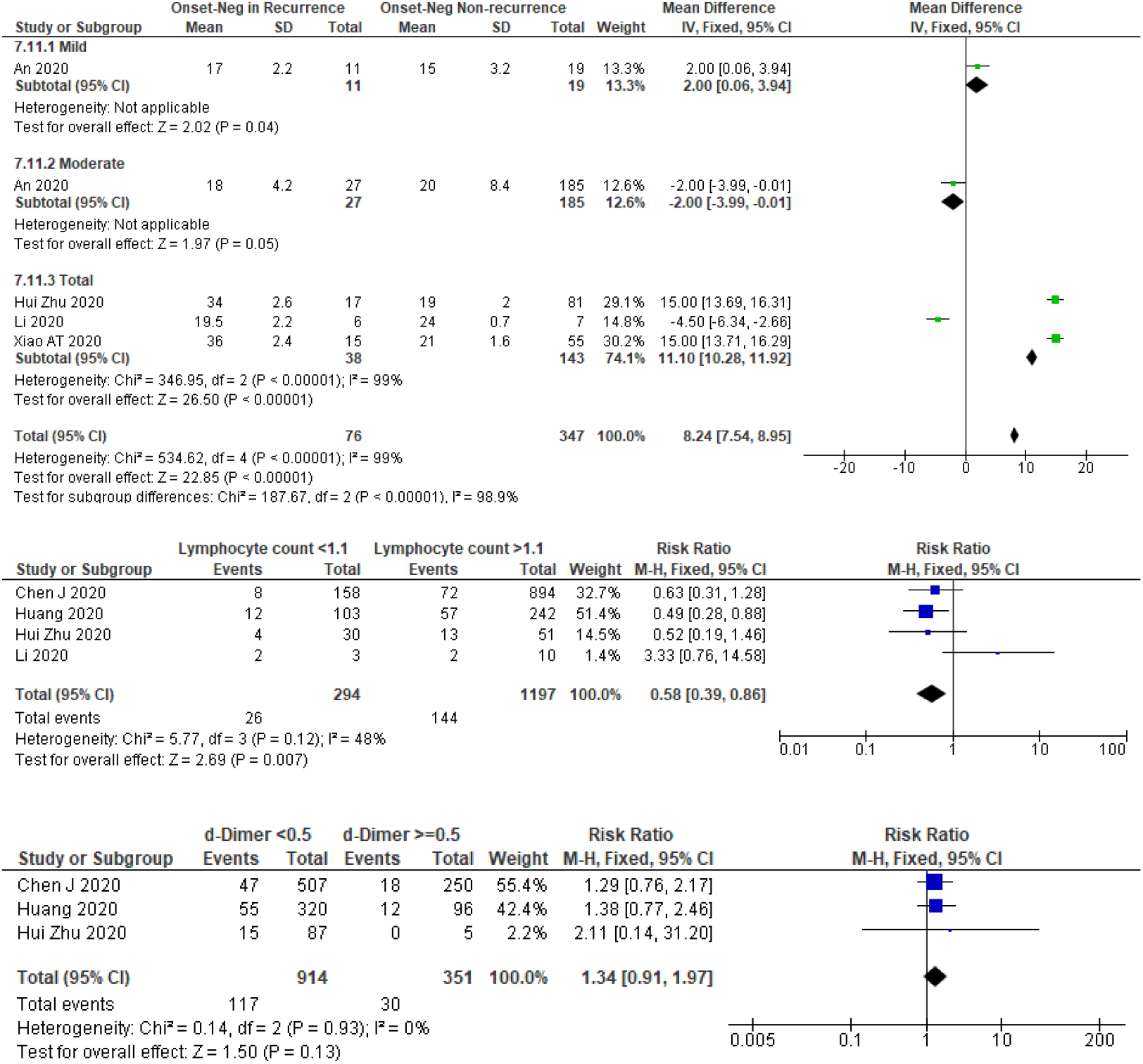
A meta-analysis of the pooled estimated RR of characteristics and clinical features to recurrent SARS-CoV-2 RNA positivity.

## Discussion

We conducted a systematic review and meta-analysis of 14 studies involving 2,568 individuals. This is the first systematic review on recurrent SARS-CoV-2 RNA positivity among individuals who have recovered from COVID-19. The pooled estimate of the incidence of recurrent SARS-CoV-2 positivity was 14.81%, confirming that recurrent positivity among patients who have recovered and been discharged from hospital is relatively common. The persistence of SARS-CoV-2 protein in some patients with recurrent SARS-CoV-2 positivity may be a sign of active viral replication and so these patients could still be infectious, although the level of infectiousness of individuals with recurrent positivity requires further evaluation. No studies in this review provided evidence of new infections in the family members or close contacts of the recovered patients that experienced recurrent positivity. Several studies clearly reported that there was no new infection infected from the patients with recurrent positivity, the study reported by Lan et al.[13] found that there we no family members infected. However, these results do not rule out the possibility that individuals with repeat positivity may still be infectious because most patients are likely to have strictly obeyed self-isolation protocols, as described by Zheng et al.[14]

We estimated that the interval between the onset of the initial episode of the disease and recurrent positivity was 35.44 days. The longest interval (50 days) was reported by Chen.[15] The time from the last negative PCR test result (used as a discharge criterion) to recurrent positivity was 9.76 days, with the longest interval (19 days) being reported by Huang.[16] Regarding the incubation period, Jing et al.[17] reported that the estimated median of incubation period was 8.13 days and the 99th percentile was 20.59 days. Considering these findings, a treatment protocol for follow-up and observation of recovered patients, should be implemented. Strict self-isolation and routine PCR retesting, where feasible, on days 9 to 20 after discharge should be considered. Routine retesting should also be considered 35 to 50 days from the onset of the illness. Further study should be conducted to elucidate whether prolonged persistent and recurrent RNA positivity still potentially infectious.

Prolonged viral shedding could be considered as the underlying mechanism of recurrent positivity as false-negative PCR test results have been reported.[18–22] The estimated duration of viral shedding based on the absence of SARS-CoV-2 RNA detection was 20 days.[23] However, the absence of nucleic acid alone cannot be used to determine whether viral shedding occurred or potential infectiousness. Viral RNA could still be detected in a long time after the disappearance of active virus,[24] and Yan et al.[25] categorized prolonged viral shedding with the cut-off of 23 days.

Our review found that younger age, a longer length of stay during the initial illness, and higher lymphocyte count was associated with an increased risk of recurrent positivity, while the presence of diabetes mellitus, severe clinical disease were associated with a reduced risk. Several studies have reported the determinants of prolonged viral shedding. A systematic review[26] concluded that the use of corticosteroid was associated with delayed viral clearing. Another review[27] reported that clearance of SARS-CoV-2 took longer in patients with gastrointestinal disease than in those with respiratory disease, especially in children. A case series reported by Huang et al. found that fecal specimens tended to have persistently detectable SARS-CoV-2 on molecular tests for longer than other specimen types.[28] Another study[25] reported that in adult patients, especially older patients had prolonged viral shedding, and that treatment with lopinavir/ritonavir was associated with a shorter shedding period. Prolonged viral shedding has also been reported to be associated with male sex, old age, concomitant hypertension, delayed admission to hospital after illness onset, severe illness at admission, invasive mechanical ventilation, and corticosteroid treatment.[29]

This review did not consider the underlying mechanism of the recurrent SARS-CoV-2 positivity; however, a previous non-systematic review[30] assessed possible mechanisms underlying recurrent SARS-CoV-2 positivity and was unable to determine whether it was attributable to false-negative results, reactivation, relapse or reinfection.

A study by Bao et al. give evidence that suggested that reinfection was unlikely. They conducted trials on Rhesus macaques that were re-infected with SARS-CoV-2 on the early recovery phase from initial infection characterized by weight loss, interstitial pneumonia, and systemic viral dissemination mainly in respiratory and gastrointestinal tracts. The results showed that primary SARS-CoV-2 infection protects from subsequent reinfection.[31] Wang et al. also reported that there is no infectious risk of COVID-19 patients with long-term fecal SARS-CoV-2 RNA positivity, and that there were no abnormalities in the gastrointestinal examination of these patients after they had been discharged.[32] However, a case report from Italy by Loconsole et al. described a case of a 48-year-old man with re-detectable positive SARS-CoV-2 after two consecutive negative SARS-CoV-2 molecular tests following his discharge from the hospital. A month after home quarantine, the man developed new symptoms of dyspnea and chest pain, causing him to re-admitted and his SARS-CoV-2 RNA test was positive on his readmission,[33] making it necessary to consider reinfection or recurrence (relapse) as possible mechanisms for recurrent SARS-CoV-2 RNA positivity on retesting. Further studies with larger sample sizes, more longer follow-up, and more detailed measurements should be conducted to determine the mechanisms underlying recurrent positivity.

Our meta-analysis produced large heterogenicity in some parameters reported; however, a sub-group analysis and meta-regression could not be conducted to identify sources of between-study heterogeneity in the pooled incidence estimates, because of insufficient study data. A further review should be re-conducted once additional publications become available.

## Conclusion

This systematic review provides evidence of SARS-CoV-2 recurrence of 14.81% among COVID-19 patients. This review also provides pooled estimated time of onset to the re-detectable positive duration was 35.44 days, and estimated time of the last negative to re-detectable positive duration was 9.76 days. Patients with younger age, no history of diabetes, mild and moderate severity, longer duration of onset to the last negative PCR, and higher lymphocyte count are more likely to experience recurrent SARS-CoV-2 positivity. Strict self-isolation for recovered patients should be well applied, and further studies should be conducted to elucidate the possibility of infectious individuals with prolonged or recurrent RNA positivity.

## Data Availability

All data generated or analyzed during this study are included in this published article or any further information could be requested to the correspondent author.

https://www.crd.york.ac.uk/prospero/display_record.php?RecordID=186306

## Abbreviations

SARS-CoV-2: Severe acute respiratory syndrome-coronavirus-2
COVID-19: coronavirus diseases-19
SMC: Semarang Medical Center
RNA: ribonucleic acid

## Author contributions

MA, AF, MR drafted the manuscript. All authors contributed to the development of selection criteria, risk of a bias assessment strategy, and data extraction criteria. MA and AF developed the search strategy, UB and RS provided statistical and methodological expertise. MR provided expertise in COVID-19 from the perspective of pulmonary medicine. UB and SA contributed to interpretation of the data. All authors read, provided feedback, and approved the final manuscript.

## Ethics approval and consent to participate

Systematic review protocol registered in PROSPERO under ID no. <u>CRD42020186306</u>

